# The Role of Prompt Engineering for Multimodal LLM Glaucoma Diagnosis

**DOI:** 10.1101/2024.10.30.24316434

**Authors:** Reem Agbareia, Mahmud Omar, Ofira Zloto, Nisha Chandala, Tania Tai, Benjamin S Glicksberg, Girish N Nadkarni, Eyal Klang

## Abstract

**Background and Aim:** This study evaluates the diagnostic performance of multimodal large language models (LLMs), GPT-4o and Claude Sonnet 3.5, in detecting glaucoma from fundus images. We specifically assess the impact of prompt engineering and the use of reference images on model performance.

**Methods:** We utilized the ACRIMA public dataset, comprising 705 labeled fundus images, and designed four prompt types, ranging from simple instructions to more refined prompts with reference images. The two model were tested across 5640 API runs, with accuracy, sensitivity, specificity, PPV, and NPV assessed through non-parametric statistical tests.

**Results:** Claude Sonnet 3.5 achieved a highest sensitivity of 94.92%, a specificity of 73.46%, and F1 score of 0.726. GPT-4o reached a highest sensitivity of 81.47%, a specificity of 50.49%, and F1 score of 0.645. The incorporation of prompt engineering and reference images improved GPT-4o’s accuracy by 39.8% and Claude Sonnet 3.5’s by 64.2%, significantly enhancing both models’ performance.

**Conclusion:** Multimodal LLMs demonstrated potential in diagnosing glaucoma, with Claude Sonnet 3.5 achieving a sensitivity of 94.92%, far exceeding the 22% sensitivity reported for primary care physicians in the literature. Prompt engineering, especially with reference images, significantly improved diagnostic performance. As LLMs become more integrated into medical practice, efficient prompt design may be key, and training doctors to use these tools effectively could enhance clinical outcomes.

## Introduction

In ophthalmology, AI has shown promise in analyzing imaging data for conditions such as age-related macular degeneration (AMD) and diabetic retinopathy and others (1–3). Recently, multi-modal large language models (LLMs) are attracting attention for their ability to process both text and visual data, which is crucial for interpreting medical images alongside clinical information (4). Such multimodal systems have the potential to improve diagnostic accuracy in various image-based tasks in ophthalmology (5).

Glaucoma, a leading cause of irreversible blindness in the western world, often progresses without symptoms until vision is severely affected (6). This makes early diagnosis crucial, but challenging. Currently, glaucoma is diagnosed through a combination of clinical exams and imaging techniques like visual field test, optical coherence tomography (OCT) and fundus photography, which captures detailed images of the retina to assess optic nerve damage (6). However, many cases go undiagnosed, with around 70% of people with glaucoma unaware they have the disease (7). Early detection is key to preventing vision loss, but the complexity of diagnosis and limited access to specialists contribute to diagnostic delays (6,7).

LLMs could offer solutions by analyzing fundus images and assisting in early glaucoma detection. With multimodal capabilities that allow them to process both text and visual data, LLMs may help reduce the diagnostic burden on ophthalmologists, allowing primary care providers to assist in earlier detection. This could shorten delays in diagnosis and accelerate treatment, especially in settings with limited access to specialists. While fundus imaging is not typically used alone for diagnosing glaucoma and often requires additional tests (6,7), showing that LLMs can reliably detect glaucoma in these images could support clinical integration. This is especially relevant as LLMs continue to advance in handling diverse types of medical data.

The current literature shows that how LLMs are prompted plays a key role in their performance (8). The wording and structure of prompts can influence how accurately the models interpret and analyze text and images. Some evidence suggests that including example images in the prompt, especially for vision-integrated tasks, improves the models’ ability to identify patterns and produce more accurate results (8,9). However, more research is needed in this area, particularly in designing field-specific prompts and guidelines for effective integration into daily clinical practice.

In this study, we aim to assess how well OpenAI’s GPT-4o and Anthropic’s Claude Sonnet 3.5 can diagnose glaucoma based on fundus images. We also examine how different prompt designs, especially those that include visual examples, affect the model’s diagnostic performance.

## Materials and Methods

### Study Design and Dataset

This study aimed to evaluate the diagnostic capabilities of two LLMs—GPT-4o and Claude Sonnet 3.5—in detecting glaucoma using fundus images. We utilized the **ACRIMA** public database (10), which comprises 705 labeled images, including 396 glaucomatous and 309 normal images (10). This dataset represents one of the largest publicly available sources for glaucoma diagnosis. The ground truth labels for the images were determined by expert ophthalmologists, providing reliable classification for training and testing purposes.

### Model Prompts and Prompt Engineering

We designed four distinct prompts to guide the LLMs in diagnosing glaucoma:

1. Simple Prompts:
  - Without Image Examples (single-shot): A basic instruction with a target image was provided to classify fundus images as “Likely Glaucomatous” or “Likely Non-Glaucomatous,” based purely on observable features in the image.
  - With Image Examples (few-shot): Along with the basic instruction and target image, we included four reference images—two glaucomatous and two non-glaucomatous—selected and validated by expert ophthalmologists. These images were intended to assist the models in their predictions.
2. Prompt-Engineered Prompts:
  - Without Image Examples (single-shot): We refined the instructions to focus on critical diagnostic features, defining the AI’s role as an expert ophthalmologist. The prompt included specific details based on the literature and expert input, such as optic nerve appearance and retinal nerve fiber characteristics.
  - With Image Examples (few-shot): This prompt combined the refined instructions and target image with the four reference images. The models were instructed to consider specific clinical markers, guided by expert advice, to improve their diagnostic accuracy.

Each prompt aimed to test whether refined instructions and the use of example images could improve the models’ diagnostic performance. Full versions of the prompts are included in the **Supplementary Materials**.

### Infrastructure

The models were implemented using Python (Ver 3.9), leveraging specific libraries for each model’s API. **GPT-4o** was accessed via the OpenAI API using the openai Python library, with model calls made through the “`client.chat.completions.create`” function. **Claude Sonnet 3.5** was accessed through the Anthropic API using the anthropic Python library, specifically the “`client.messages.create`” function.

Both models were provided with asbase64-encoded images, as required by their respective APIs. Each image in the dataset was classified four times by both models: once with the simple prompt, once with the simple prompt containing examples, once with the prompt-engineered version, and once with the prompt-engineered version containing examples. For each classification, the models were instructed to return a binary result: 0 for “Likely Non-Glaucomatous” and 1 for “Likely Glaucomatous.” This process resulted in a total of 5640 API runs across both models (**Figure 1**).

**Figure 1:**
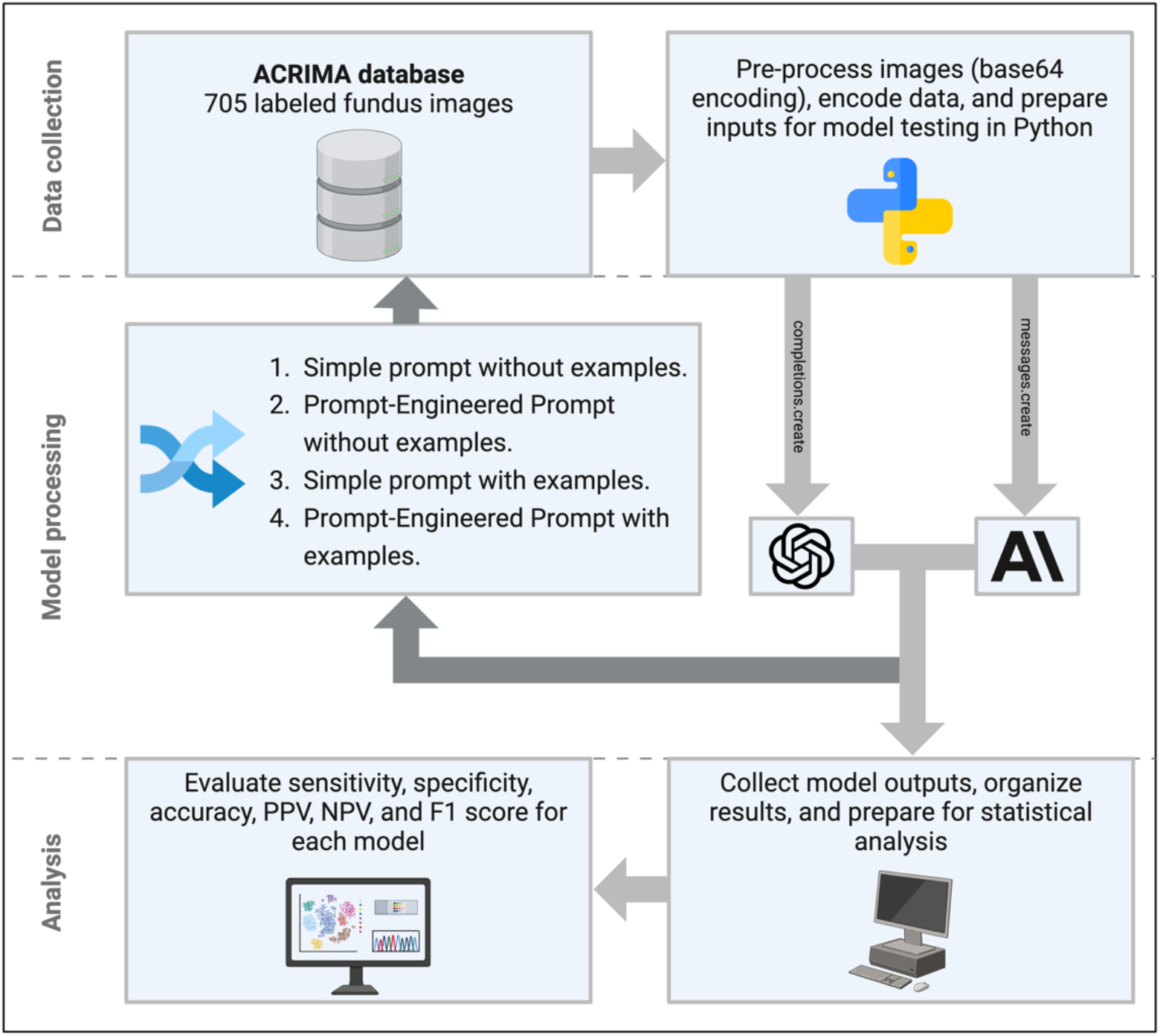
A flowchart of the study design.

### Statistical Analysis

Statistical analysis assessed the diagnostic performance of each model iteration using descriptive metrics—accuracy, sensitivity, specificity, positive predictive value (PPV), and negative predictive value (NPV). Data normality was tested with the Shapiro-Wilk test, and due to non-normal distributions, non-parametric tests were used. The Kruskal-Wallis test identified significant differences across model iterations, and pairwise comparisons were made using the Mann-Whitney U test. Levene’s test assessed variance homogeneity. F1 score for overall performance were calculated, and the Mann-Whitney U test compared GPT-4o and Claude Sonnet 3.5 across glaucoma and non-glaucoma datasets. Detailed statistical methods are provided in the **Supplementary Materials**.

## Results

### Overall Model Performance

The diagnostic performance of GPT-4o and Claude Sonnet 3.5 was evaluated across four prompt iterations. For confirmed glaucoma cases, the mean correct diagnoses ranged from 46.95% to 94.92%. GPT-4o’s correct diagnoses rates ranged from 62.44% to 81.47% across its iterations, while Claude Sonnet 3.5 showed higher performance, with correct diagnoses rates ranging from 46.95% to 94.92%. In non-glaucoma cases, the mean correct diagnoses percentage varied between 26.54% and 79.29%, with GPT-4o performing in the range of 49.51% to 79.29% and Claude Sonnet 3.5 ranging from 26.54% to 73.46%.

Regarding diagnostic statistics, sensitivity ranged from 46.95% to 94.92%, while specificity ranged from 20.71% to 73.46% across the models and iterations. Accuracy varied between 48.51% and 85.49%. PPV ranged from 53.07% to 82.02%, and NPV showed similar variability, spanning 35.36% to 91.90%. **Table 1** summarizes the results for each iteration.

**Table 1.**
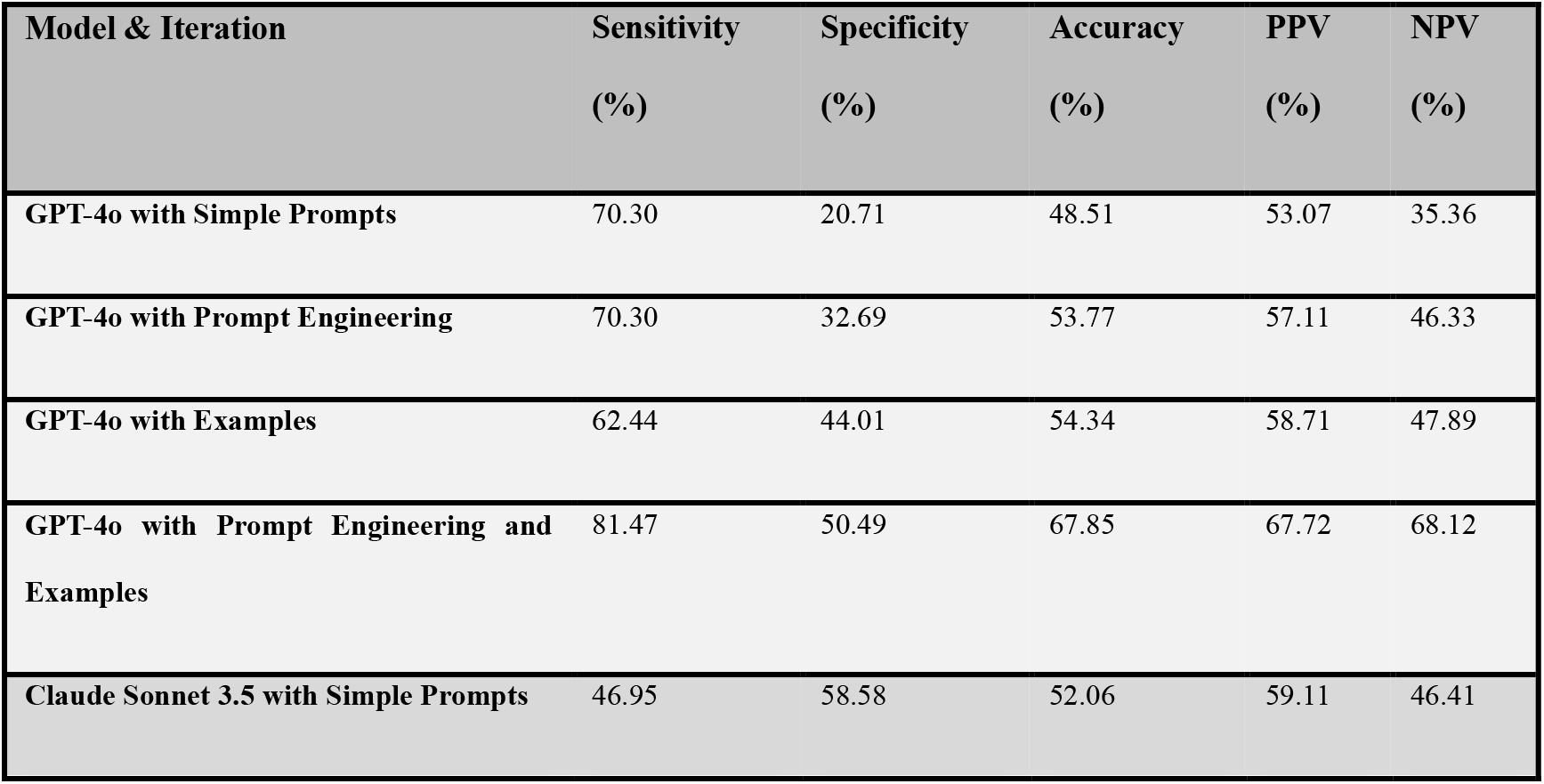
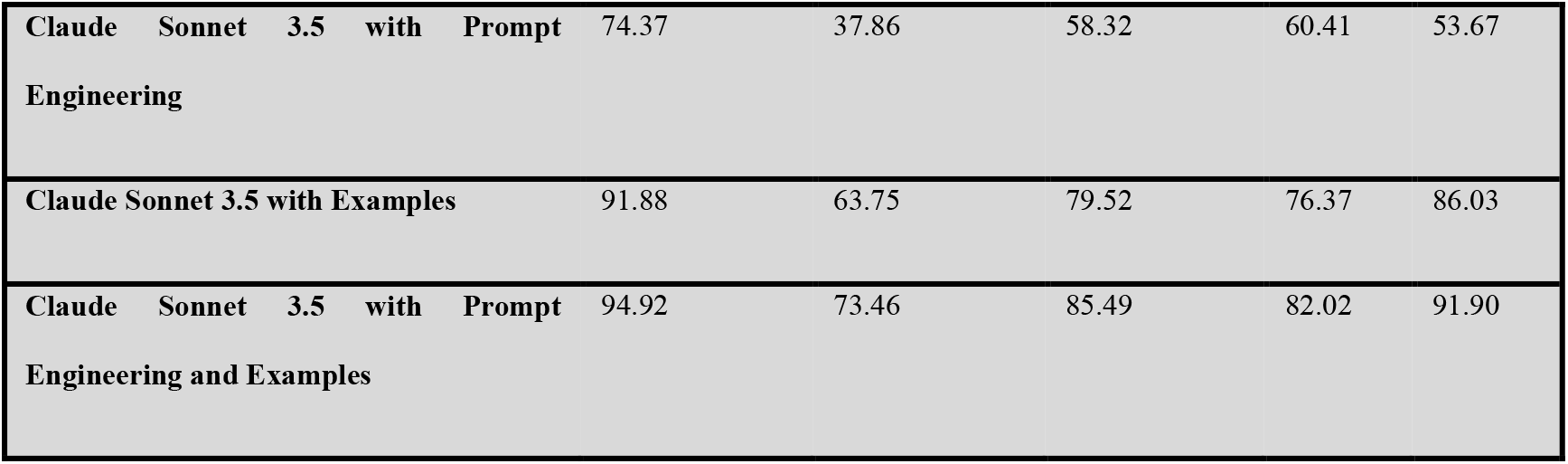
Diagnostic Statistics for Each Model Iteration.

| Model & Iteration | Sensitivity (%) | Specificity (%) | Accuracy (%) | PPV (%) | NPV (%) |
| --- | --- | --- | --- | --- | --- |
| GPT-4o with Simple Prompts | 70.30 | 20.71 | 48.51 | 53.07 | 35.36 |
| GPT-4o with Prompt Engineering | 70.30 | 32.69 | 53.77 | 57.11 | 46.33 |
| GPT-4o with Examples | 62.44 | 44.01 | 54.34 | 58.71 | 47.89 |
| GPT-4o with Prompt Engineering and Examples | 81.47 | 50.49 | 67.85 | 67.72 | 68.12 |
| Claude Sonnet 3.5 with Simple Prompts | 46.95 | 58.58 | 52.06 | 59.11 | 46.41 |
| Claude Sonnet 3.5 with Prompt Engineering | 74.37 | 37.86 | 58.32 | 60.41 | 53.67 |
| Claude Sonnet 3.5 with Examples | 91.88 | 63.75 | 79.52 | 76.37 | 86.03 |
| Claude Sonnet 3.5 with Prompt Engineering and Examples | 94.92 | 73.46 | 85.49 | 82.02 | 91.90 |

### Comparison of Models and Iterations

A significant improvement in performance was observed across both GPT-4o and Claude Sonnet 3.5 when both prompt engineering and example images were incorporated.

For GPT-4o, sensitivity increased from 70.30% with simple prompts to 81.47% with prompt engineering and examples (p < 0.001). Similarly, accuracy improved from 48.51% to 67.85%, and PPV increased from 53.07% to 67.72% (p < 0.001 for both). Specificity, though lower for initial prompts (20.71%), reached 50.49% when both prompt engineering and examples were used.

For Claude Sonnet 3.5, the improvement was more pronounced. Sensitivity increased from 46.95% with simple prompts to 94.92% with prompt engineering and examples (p < 0.001). Accuracy also rose from 52.06% to 85.49%, and PPV improved from 59.11% to 82.02% (p < 0.001). Specificity, which started at 58.58%, increased to 73.46% in the same iteration.

Pairwise Wilcoxon rank-sum tests revealed statistically significant differences between model iterations, showing that prompt engineering and the inclusion of examples consistently enhanced model performance (p < 0.001 across iterations). **Figure 2** highlights these improvements.

**Figure 2:**
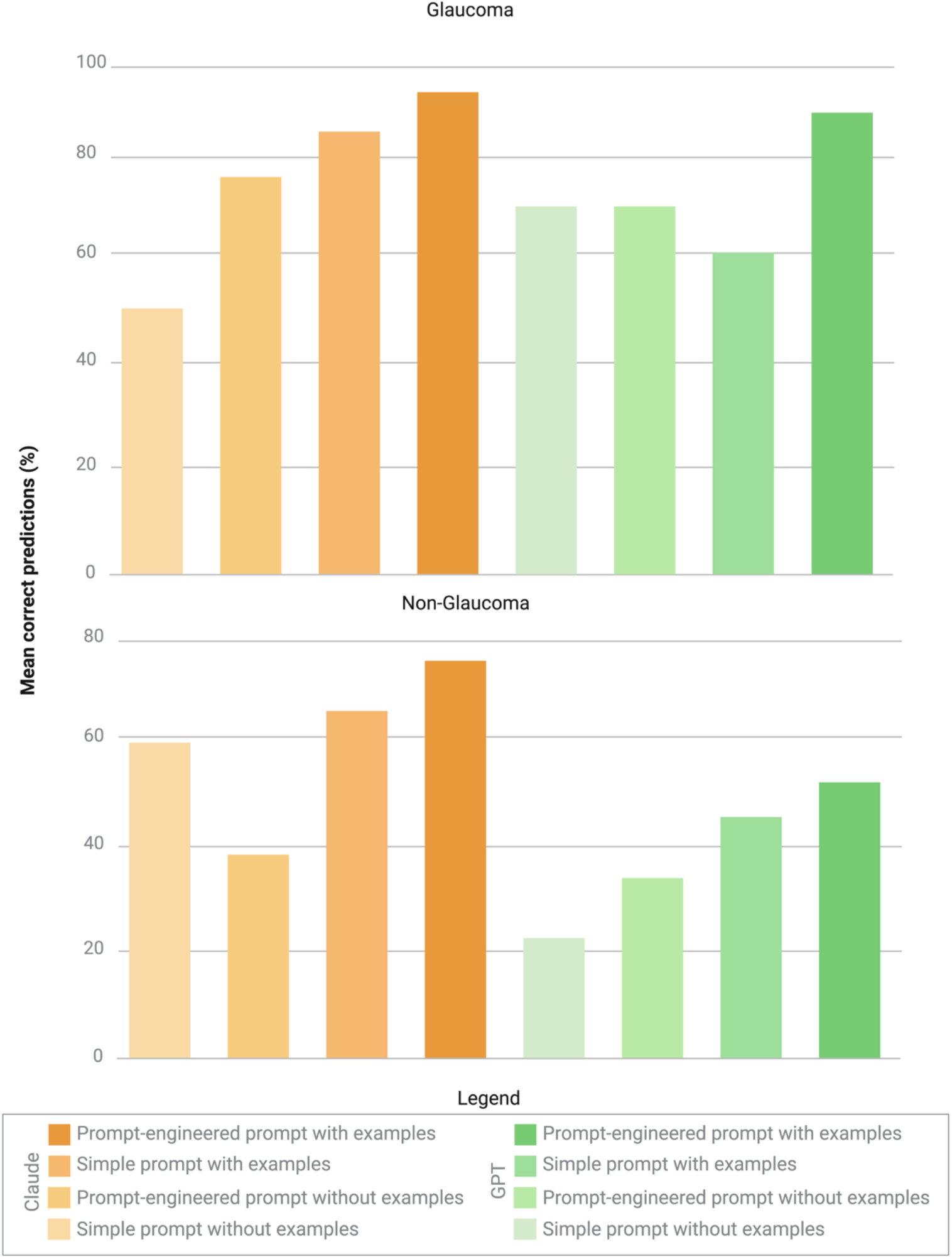
Correct prediction by model and prompt type in glaucomatous vs non-glaucomatous cases.

### Overall Comparison

The overall performance of GPT-4o and Claude Sonnet 3.5 was compared using the F1 score, which balances precision and recall. The Mann-Whitney U test showed no statistically significant difference between the two models (p = 0.485). The mean F1 score for GPT-4o across all iterations was 0.645, while Claude Sonnet 3.5 achieved a mean F1 score of 0.726.

## Discussion

This study evaluates the diagnostic capabilities of two multimodal LLMs: GPT-4o and Claude Sonnet 3.5, in detecting glaucoma using fundus images, focusing on how different prompt designs influence their performance. The results indicate that incorporating prompt engineering and reference images significantly improved both models’ diagnostic accuracy. Notably, Claude Sonnet 3.5 outperformed GPT-4o in overall sensitivity and specificity, especially in iterations using both prompt engineering and visual examples.

GPT-4o and Claude Sonnet 3.5 demonstrated significant potential in diagnosing glaucoma, particularly when using prompt engineering and reference images. When compared to the performance of primary care physicians, LLMs performed notably better. For instance, Wada-Koike et al. reported that non-ophthalmologists, including primary care physicians, achieved a sensitivity of 22% and a specificity of 92% when interpreting fundus images for glaucoma screening (11). In contrast, Claude Sonnet 3.5 reached a sensitivity of 94.92% and specificity of 73.46% with prompt engineering, showing a marked improvement in detecting glaucoma cases. While the LLM’s specificity was lower than that of primary care physicians, its much higher sensitivity indicates the potential to catch more true glaucoma cases earlier, which is especially important in primary care settings where access to specialists may be limited.

When comparing our LLMs to ophthalmology residents and experts, Claude Sonnet 3.5 achieved a sensitivity of 94.92% and an accuracy of 85.49%, surpassing the upper sensitivity range of residents (12), whose sensitivity ranged from 72.6% to 91.2% and specificity from 72.8% to 82.4% (12). This also aligns closely with the diagnostic accuracy of experienced ophthalmologists, who typically achieve around 80.5% accuracy in classifying optic disc photographs for glaucoma, with sensitivities exceeding 85% (13). However, when compared to deep learning convolutional neural networks (CNNs) models, LLMs still fall short in overall balanced performance. For instance, Phene et al. reported a CNN algorithm with an AUROC of 0.945 (14), and CNN systems developed by Ahn et al. and Li et al. achieved AUROCs of 0.94 and 0.986, respectively, with sensitivities of 95.6% and specificities over 90% (15,16). While the sensitivity of Claude Sonnet 3.5 is competitive with these models, its lower specificity (73.46%) indicates that further refinement is required for LLMs to match the more balanced accuracy of deep learning systems.

In comparison to a similar study by AlRyalat et al., where GPT-4 achieved 90% accuracy, 50% sensitivity, and 94.44% specificity using the REFUGE dataset (17), our GPT-4o showed a lower accuracy, ranging from 48.51% to 67.85%, but higher sensitivity, ranging from 62.44% to 81.47%, though with lower specificity, ranging from 20.71% to 50.49%, across different prompt iterations. The differences may stem from our use of the larger ACRIMA dataset with 705 images and our focus on prompt engineering, which included detailed instructions and reference images. This approach may have improved sensitivity but reduced specificity, as the more complex prompts helped in identifying glaucomatous cases while making it harder to accurately classify non-glaucomatous ones. In another a related study using GPT-4 Vision Preview on 100 fundus images from the ACRIMA dataset, GPT-4 achieved an accuracy of 68.2% and a sensitivity of 70.7% (18). Our results showed similar sensitivity, but with greater variability due to the larger dataset and the use of both GPT-4o and Claude Sonnet 3.5 models. While GPT-4o’s accuracy was slightly lower, ranging from 48.51% to 67.85%, we specifically evaluated the quantitative impact of prompt engineering across two models, further highlighting the effect of detailed prompts and visual examples on diagnostic performance.

However, LLMs present several advantages over CNNs, particularly in their ability to handle both clinical and imaging data, offering a more comprehensive diagnostic tool (19). For instance, deep learning models require more retraining and Graphics processing unit (GPU) resources (20), which may not be accessible in all clinical environments. In contrast, LLMs like GPT-4o and Claude Sonnet 3.5 do not require extensive retraining and can be integrated more easily into primary care settings. This is particularly important for early glaucoma detection in non-specialist environments, where there is limited access to ophthalmologists. Given that close to 70% of glaucoma cases remain undiagnosed (7), LLMs could serve as an accessible first-line diagnostic tool to bridge this gap. The relatively lower computational demand of LLMs also makes them a more feasible option in resource-constrained settings, potentially allowing for early intervention in areas with limited medical infrastructure. This capacity to integrate multimodal data (text and imaging) and function with less computational power underscores the practical promise of LLMs, despite their current diagnostic limitations compared to more specialized deep learning systems.

This study has limitations. First, the models were not fine-tuned for glaucoma detection, which likely impacted their overall accuracy. While prompt engineering improved their performance, they still did not reach the precision of established deep learning systems. Additionally, the study relied solely on fundus images, whereas glaucoma diagnosis typically involves multiple clinical tests like visual field test, OCT etc., limiting the models’ applicability in real-world settings. A study should be conducted to assess the capability of LLMs in identifying glaucoma through visual field tests and OCT.The use of open-access data, which may overlap with the models’ training data, also raises concerns about potential bias. Although LLMs learn patterns rather than memorizing specific examples, this overlap could affect the generalizability of the results. Future studies should validate these models on independent datasets and incorporate multimodal data, including clinical records, for more comprehensive and reliable diagnostic capabilities.

*In conclusion*, this study demonstrates the potential of multimodal LLMs in diagnosing glaucoma from fundus images, with significant improvements in performance through prompt engineering. The models showed sensitivity comparable to CNNs and exceeded the diagnostic performance of primary care physicians and ophthalmology residents in some cases. While LLMs do not yet fully match the accuracy of CNNs, they offer advantages like ease of use, lower computational demands, and the ability to process multimodal data, making them potentially valuable for early glaucoma detection.

## Data Availability

All data produced in the present study are available upon reasonable request to the authors

