## Supplementary Materials for "The Role of Prompt Engineering for Multimodal LLM Glaucoma Diagnosis"

##### Evaluating Multimodal LLMs in Glaucoma Diagnosis: The Role of Prompt Engineering

Mount Sinai

Division of Data-Driven and Digital Medicine (D3M), Icahn School of Medicine at Mount Sinai, New York, USA

### Section 1: The prompts

- **Simple prompt without examples:**

Hello, your task is to perform a preliminary analysis of the attached fundus photograph to determine whether they show signs of Glaucoma. You are required to classify the photograph as either 'Likely Glaucomatous' or 'Likely Non-Glaucomatous' based on observable features.

Classify this image as either 'Likely Glaucomatous' (1) or 'Likely Non-Glaucomatous' (0). Respond with only 0 or 1.

- **Prompt-Engineered Prompt without examples:**

You are an expert ophthalmologist specializing in glaucoma detection through the analysis of fundus photographs. Your task is to assess the attached fundus photograph and provide a preliminary diagnosis, classifying it as either "**Likely Glaucomatous**" or "**Likely Non-Glaucomatous**".

When analyzing the image, focus on key indicators such as:

- Optic disc cupping
- Neuroretinal rim thinning
- Peripapillary atrophy
- Retinal nerve fiber layer defects

Ensure the diagnosis is balanced, avoiding both overdiagnosis and underdiagnosis. Your goal is to offer a well-reasoned, expert-level judgment based on observable features.

Classify this image as either 'Likely Glaucomatous' (1) or 'Likely Non-Glaucomatous' (0). Respond with only 0 or 1.

- **Simple prompt with examples:**

Hello, your task is to perform a preliminary analysis of the attached fundus photograph to determine whether they show signs of Glaucoma. You are required to classify the photograph as either 'Likely Glaucomatous' or 'Likely Non-Glaucomatous' based on observable features.

Classify this image as either 'Likely Glaucomatous' (1) or 'Likely Non-Glaucomatous' (0). Respond with only 0 or 1. Use the provided example images as reference templates for accurate classification.

- **Prompt-Engineered Prompt with examples:**

You are an expert ophthalmologist specializing in glaucoma detection through the analysis of fundus photographs. Your task is to assess the attached fundus photograph and provide a preliminary diagnosis, classifying it as either **"Likely Glaucomatous"** or **"Likely Non-Glaucomatous"**.

When analyzing the image, focus on key indicators such as:

- Optic disc cupping
- Neuroretinal rim thinning
- Peripapillary atrophy
- Retinal nerve fiber layer defects

Ensure the diagnosis is balanced, avoiding both overdiagnosis and underdiagnosis. Your goal is to offer a well-reasoned, expert-level judgment based on observable features.

Classify this image as either 'Likely Glaucomatous' (1) or 'Likely Non-Glaucomatous' (0). Respond with only 0 or 1. Use the provided example images as reference templates for accurate classification.

#### Section 2: Validated image references for model learning

##### Unlikely Glaucomatous two expert ophthalmologist validated images:

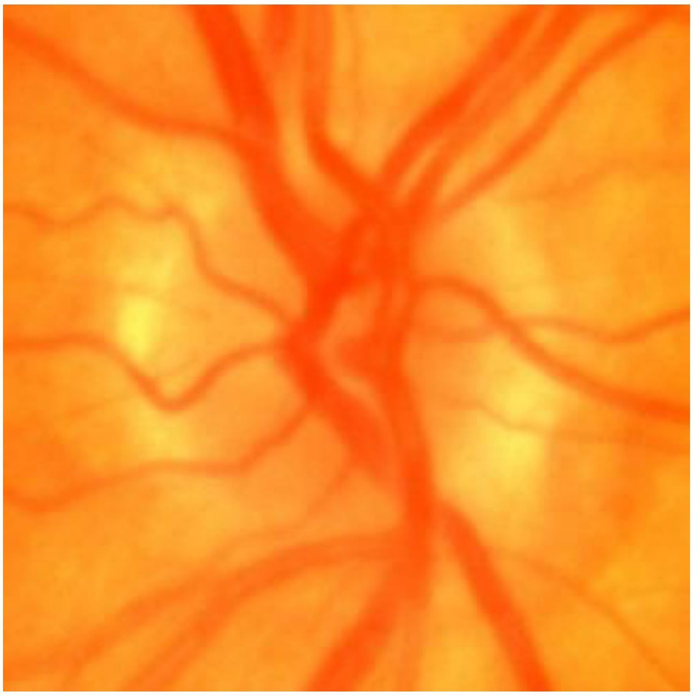

Unlikely Glaucomatous reference image 1

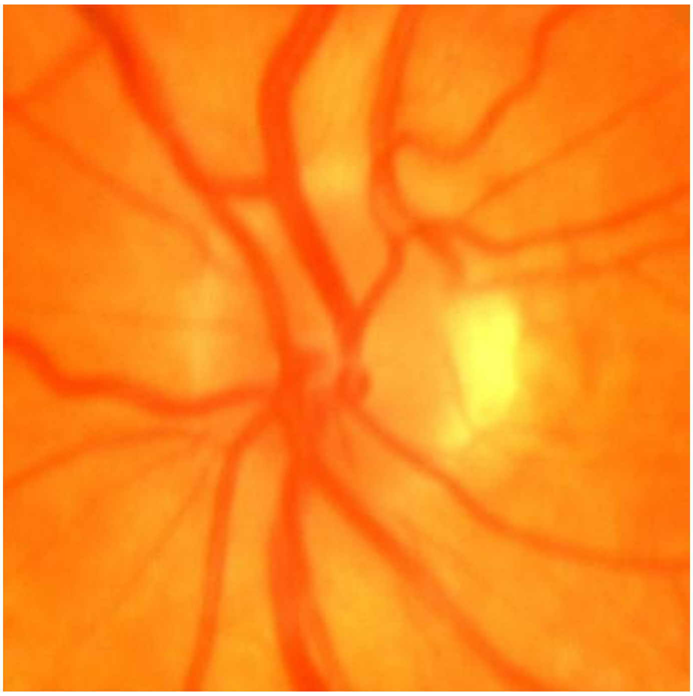

Unlikely Glaucomatous reference image 2

#### Likely Glaucomatous two expert ophthalmologist validated images:

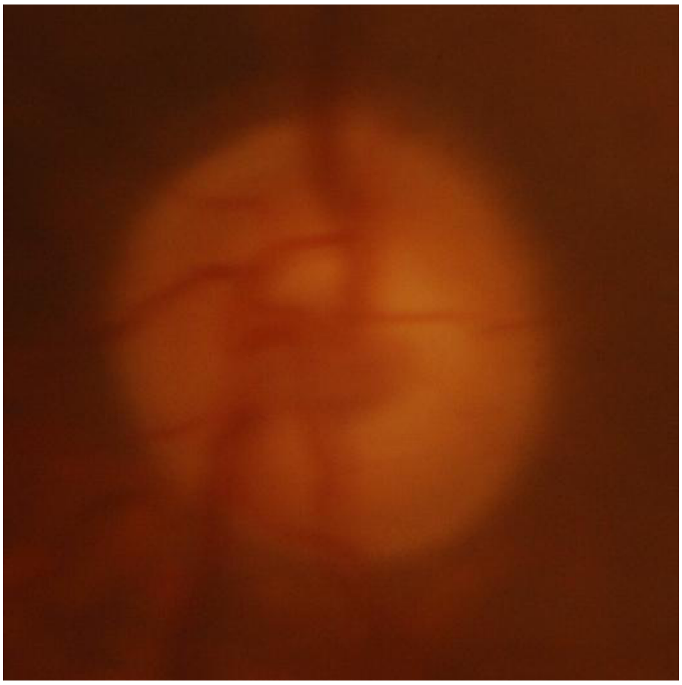

Likely Glaucomatous reference image 1

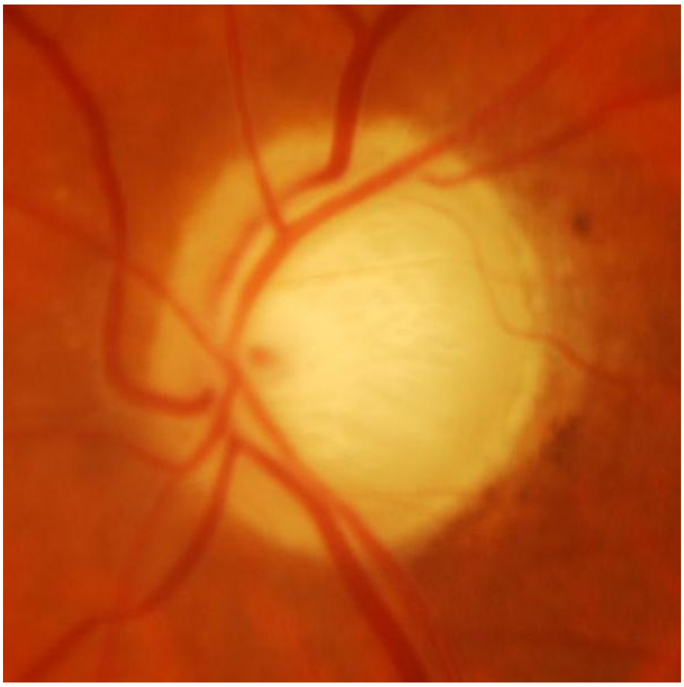

Likely Glaucomatous reference image 2

### Section 3: Detailed Statistical Analysis

In this section, we outline the detailed statistical methodology used to evaluate and compare the performance of GPT-4o and Claude Sonnet 3.5 for glaucoma detection.

**1. Descriptive Statistics**

For each model iteration, we calculated descriptive statistics, including:

- **Mean** and **Standard Deviation (SD)** for accuracy, sensitivity, specificity, positive predictive value (PPV), and negative predictive value (NPV). These were computed across all test images, both glaucoma and non-glaucoma, for all four prompt types (simple, simple with examples, prompt-engineered, and prompt-engineered with examples).

**2. Normality Testing**

We first tested the normality of the distribution for each performance metric using the **Shapiro-Wilk Test**:

- For each model iteration and each diagnostic metric, the test was applied to determine whether the data followed a normal distribution.
- **Result**: Most data were not normally distributed (Shapiro-Wilk p-value < 0.05), leading to the use of non-parametric statistical tests.

**3. Non-Parametric Tests**

Given the non-normal distribution of the data, non-parametric statistical methods were applied:

- **Kruskal-Wallis Test**:
    - Used to assess whether there were statistically significant differences in performance between the four model iterations for each model (GPT-4o and Claude Sonnet 3.5).
    - Applied separately for the glaucoma and non-glaucoma datasets.
    - Null Hypothesis (H0): All model iterations have the same performance distribution.
    - **Interpretation**: A p-value < 0.05 indicated significant differences between the model iterations.
- **Mann-Whitney U Test** (Wilcoxon rank sum test):

- Pairwise comparisons were conducted between individual model iterations within each dataset (e.g., comparing GPT_WE to GPT++, Claude_WE to Claude++).
- Additionally, GPT-4o and Claude Sonnet 3.5 were compared to assess whether there were significant differences in their overall performance across all iterations.
- **Bonferroni correction** was applied to adjust p-values for multiple comparisons, ensuring conservative estimates of significance.

**4. Variance Testing**

- **Levene's Test** was used to assess the homogeneity of variances across model iterations, ensuring that the data met the assumptions required for the applied tests.
  - Null Hypothesis (H0): The variances between the model iterations are equal.
  - A p-value < 0.05 indicated that the variances were significantly different, suggesting variability in performance consistency across iterations.

**5. F1 Score**

- The F1 Score was calculated for each model by combining the precision (PPV) and recall (sensitivity) across all prompt types and iterations to provide a balanced performance measure
  - The overall performance of GPT-4o and Claude Sonnet 3.5 was compared using the F1 scores.
  - The Mann-Whitney U Test was used to compare the F1 scores between GPT-4o and Claude Sonnet 3.5 to quantify their general diagnostic performance across both glaucoma and non-glaucoma datasets.

**6. Sensitivity, Specificity, PPV, and NPV Calculations**

We calculated the sensitivity, specificity, PPV, and NPV for each model iteration using the following formulas:

- **Sensitivity**: True Positives / (True Positives + False Negatives)
- **Specificity**: True Negatives / (True Negatives + False Positives)
- **PPV (Positive Predictive Value)**: True Positives / (True Positives + False Positives)

- **NPV (Negative Predictive Value)**: True Negatives / (True Negatives + False Negatives)

Each of these values was calculated separately for both glaucoma and non-glaucoma datasets, and the results were averaged across all prompt types and iterations to provide a holistic performance view.
